# The impact of income-support interventions on life course risk factors and health outcomes during childhood: a systematic review in high income countries

**DOI:** 10.1101/2022.09.16.22280026

**Authors:** Delia Boccia, Silvia Maritano, Costanza Pizzi, Matteo G Richiardi, Sandrine Lioret, Lorenzo Richiardi

**Affiliations:** Faculty of Population and Health Policy, London School of Hygiene and Tropical Medicine, 15-17 Tavistock Pl, London WC1H 9SH, United Kingdom; Department of Medical Sciences, University of Turin and CPO-Piemonte, Turin, Italy; University School for Advanced Studies IUSS Pavia, Italy; Centre for Microsimulation and Policy Analysis, Institute for Social and Economic Research, University of Essex, Colchester, UK; Université Paris Cité, INSERM, INRAE, CRESS, Paris, France

## Abstract

In high income countries one in five children still lives in poverty. This is known to adversely shape the life course health trajectory of these children; however, much less is understood on whether social and fiscal policies have the capacity to reverse this damage, which intervention is likely to be most effective and when these interventions should be delivered to maximise their impact. This systematic review attempts to address these questions by looking at the impact of income-support interventions delivered during the first 1,000 days of life on cardiovascular, metabolic, respiratory and mental health outcomes. The review was restricted to experimental or quasi experimental studies conducted in high income countries. Studies of interest were retrieved from multidisciplinary database as well as health, economic, social sciences-specific literature browsers. Evidence of interest were summarised via narrative synthesis approach. Robustness of findings was assessed by tabulating impact by health outcome, type of intervention and study design. Overall, 18 relevant papers were identified, including 16 independent studies, one meta-analysis of randomized control trials (RCTs) and one pooled analysis of RCTs. Income-support interventions included: unconditional/conditional cash transfers, income tax credit, welfare to work, and minimum wage salary policies. Most studies were conducted in North America. Overall, the evidence suggested a positive, albeit small, effect of most policies on birth weight outcomes, but limited effect on mental health indicators. Results seemed to be robust to the type of intervention, but not to the study design, with RCTs consistently less likely to detect an impact. Given the large number of people targeted by these programs, one could infer that – despite small – the observed effect may be still relevant at population level. Nonetheless, the limited generalisability of the evidence gathered hampers firm conclusions. For the future, the breath and scope of this literature need to be broadened to fully exploit the potential of these interventions and understand how their public health impact can be maximised.

## Introduction

### Background

Despite the overall global improvement of most development indices, one in five children in high income countries still lives in poverty, with striking variation across countries in terms of prevalence.(1) In one recent analysis from UNICEF involving 41 high income countries Denmark showed the best record on relative poverty, but even in such affluent country 9.2 per cent of children are considered poor (defined as living in a household with income below 60% of the median household after housing costs). Israel and Romania showed the worst records on relative poverty, with more than one child in three falling below the poverty line. Bulgaria, Mexico, Spain, Turkey and the United States also have child poverty rates substantially greater than the rich-world average.(1) Recently, some high income countries are witnessing a rise in childhood poverty: in the United Kingdom, for example, child poverty rose by two percentage points between 2014 and 2017, and it is projected to increase further through to 2022.(2) These forecasts are likely to having been exacerbated by the Covid-19 pandemic.(3) A mounting body of evidence suggests unequivocally that exposure to adverse socioeconomic circumstances during foetal life and early childhood affects clinical, behavioural and cognitive outcomes and - most importantly - can shape later life health trajectories.(4) These socioeconomic inequalities are preventable and unfair, particularly in the case of children who have little control over their health and the factors that influence it.(5)

Overall, strategies to prevent, reduce and mitigate child poverty and its consequences generally involve three key components—support of early childhood education and care, income redistribution through cash and or in-kind benefits and tax systems, and policies to increase the employment chances and wages of families living in poverty.(6, 7) These measures are considered to play a crucial role in reducing child inequalities mainly by increasing children’s human capital, reducing their vulnerability to the financial and physical consequences of ill-health and overall by interrupting the intergenerational transmission of poverty.

While there is evidence that all three components are likely to be effective at reducing child poverty globally,(6) at least in high income countries, few experimental and quasi-experimental studies have sought to determine whether the poverty effect of these macro-level interventions translate into a positive child-health effect,(8) Even less is known about whether some approaches are more likely to lead to greater health benefits than others, and whom and when is likely to most benefit from these interventions.

Important knowledge gaps remain also in terms of: a) *how* socioeconomic disadvantage experienced during early childhood *biologically* affects individuals’ life course health trajectories; and b) the extent to which the biological damages are exerted by socioeconomic disadvantage and *how* these biological damages can be effectively prevented and/or repaired through interventions able to address income inequalities during the first 1,000 days of life (from pregnancy to age 2).

This review aims to contribute to these knowledge gaps by providing an evidence synthesis of the child health impact of macro-level socioeconomic interventions, and in particular of income support policies, delivered in the 1,000 days of life. This effort is part of the LifeCycle project, an EU Horizon 2020-funded (2017-2022) project, whose scope is to leverage knowledge from a network of EU child cohorts in order to: 1) identify markers of early-life stressors affecting health throughout the lifecourse, including socioeconomic, lifestyle, migration and urban environment ones, and 2) translate the findings into policy recommendations for targeted prevention strategies.(9)

### The conceptual framework

Early life socioeconomic stressors can affect life course cardiometabolic, respiratory and mental health outcomes through epigenetic mechanisms, fetal and childhood development and adaptation, and finally by influencing the differential burden of life course risk factors and health outcomes during early life. In order to identify entry points for interventions, this framework needs to be further unpacked to elucidate the pathways through which socioeconomic disadvantage arises, operates and is perpetuated **(Figure 1)**.

**Figure 1.**
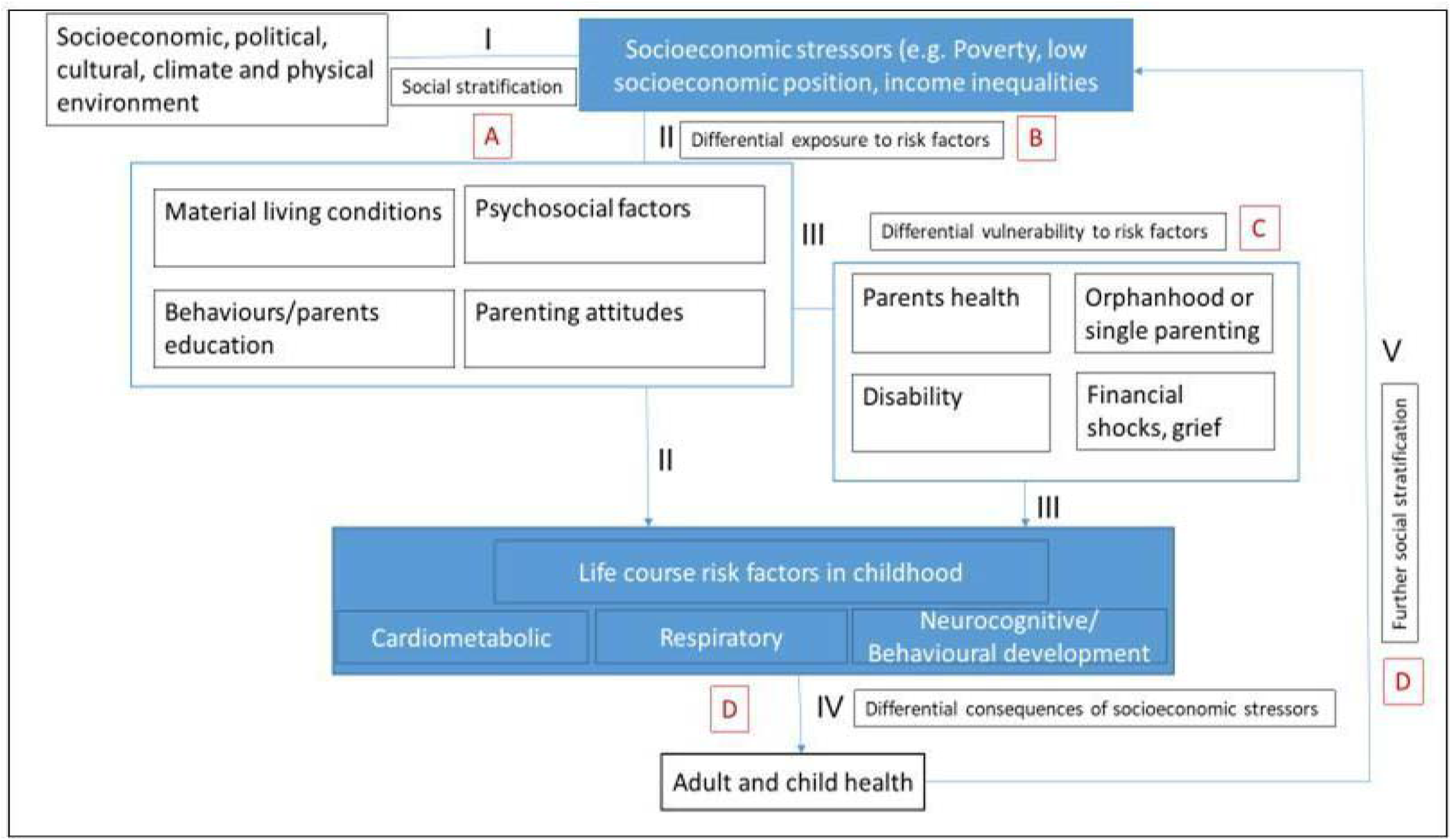
LifeCycle conceptual framework: impact of socio-economic stressors on life course risk factors.

Following from Diderichsen and colleagues conceptual model,(5) we can assume that the primer drivers of socioeconomic stressors in childhood are positioned at distal level and refer to those structures and constructs that influence the socioeconomic position of individuals in a society ***(Fig 1, Pathway I)***. Socioeconomic differences can influence the *differential exposure* to important material, psychosocial and behavioral risk factors ***(Fig 1, Pathway II)*** or can affect the *differential susceptibility* of children to these risks (***Fig 1, Pathway III***, e.g. the impact of any given risk factor may be more pronounced in less advantaged groups due to their greater likelihood of being exposed to other important and interacting risk factors). Finally, socioeconomic stressors may influence the *differential vulnerability* of children to the clinical and financial consequences of health conditions during childhood ***(Fig 1, Pathway IV)***, which ultimately can further exacerbate the disadvantage in early life and adulthood ***(Fig 1, Pathway V)***.

Depending on the pathway we can identify different entry points for interventions as outlined in **Table 1**. For the purpose of this review, and consistent with the objectives of LifeCycle, we decided to concentrate on distal-level interventions that directly affect social stratification, and are aimed at reducing inequalities through educational, labor market, welfare and poverty alleviation strategies (**Pathway I, Intervention A**). Within this broad group of interventions, we focussed on income support interventions that are defined as all measures taken by authorities aimed at providing an adequate income to their citizens via different benefit schemes, which are implemented within different policies with different aims and objectives.(10) Their implementation may embrace different criteria of selectivity and generosity across each intervention and setting. Overall, income support programs are hypothesized to improve child and adolescent outcomes via the family investment model, according to which families have more money to spend on inputs(11, 12) or more time to spend with children,(13) and the family stress model, according to which maternal depression and stress are lower because household resources are higher.(14)

**Table 1.** Examples of entry points for interventions that address socioeconomic stressors in the early life.

| **Pathway** | **Interventions** |
| --- | --- |
| *I. Creation of social inequalities and disadvantage* | **A.** Policies that influence the process of social stratification through educational system, labour market, taxation and legislation, welfare and poverty-alleviation strategies. |
| *II. Differential exposure to risk factors* | **B.** As above, but also policies that include classic public health interventions that improve housing, working conditions, and access to education and health services |
| *III. Increased vulnerability to risk factors* | **C.** Policies that include both social and public health intervention in a multisectorial/coordinated fashion to address the amplified health impact among children experiencing multiple risk factors at the same time. |

### Why this review is important

Recently, at least three systematic reviews have attempted to explore the child health impact of poverty alleviation strategies.(15-17)

They differ from our own work in terms of scope and key inclusion criteria. The systematic review and meta-analysis from Courtin et al(15) included a wide range of social policies/poverty alleviation strategies including housing, education and health insurance-related interventions, examined only RCTs, and looked at the impact on health in the general population rather than just on child’s health only.

Another recent review, from Cooper et al,(16) examined the effect of household income itself rather than socioeconomic interventions able to modify income and included as exposure of interest also lotteries and income shocks. They did not restrict their inclusion criteria to RCTs and quasi-experimental studies, but also included findings from observational studies and extended their focus beyond strict health outcomes, including school achievement.

Finally, Simpson et al(17) focused on the impact of social security benefits - more precisely the effect of changes in the eligibility and the amount and type of benefits provided -on a number of mental health indicators both in adults and children. Only observational studies were included in this review.

Our review adds to this body of knowledge by setting more stringent inclusion criteria and restricting its boundaries of investigation. Specifically, this review aimed to generate evidence on the impact of interventions able to modify the effect of early-life socioeconomic stressors during the first 1,000 days of life; and in order to more specifically link and interpret our findings according to potential underlying mechanisms, we focused on those interventions that affected income inequalities. While the results of previous studies suggested some effects of socioeconomic policies on overall physical and mental health outcomes, we aimed to assess whether these interventions can affect children specific cardiovascular, metabolic, respiratory and mental health outcomes, relying only on results coming from either experimental or quasi experimental studies.

## Material and Methods

The methods used in this review largely followed the recommendations of Waddington et al on the review of international development interventions.(18) With the exception of the search strategy definition and roll out, all steps were undertaken in parallel from at least two authors of this report.

### Search strategy and databases

Electronic searches have covered key bibliographic databases including:

⍰ Multidisciplinary ones, such as SCOPUS, Web of Science and Google Scholar;
⍰ Specific to social sciences, both general and discipline-specific, such as Social Science Research Network (SSRN), and Econlit for economics, PsycInfo for behavioural studies;
⍰ Specific to biomedical research, including Pubmed/Medline, EMBASE;
⍰ The Cochrane Library Central for both trials and reviews registry.

Consistent with existing recommendations,(18) we adopted a ‘snowballing’ approach: starting from important primary studies and already existing reviews we further increased the body of references both by bibliographic back-referencing and citation tracking (i.e. reviewing references in which the included study has been cited).

In terms of search strategy, we focused on two groups of key terms to begin with:

**GROUP 1**- Social welfare OR Social protection OR Cash/food/in-kind transfers OR child grants OR child benefits OR child allowances OR Tax benefits OR Child tax credit or Work-based programmes;

**GROUP 2** – child health.

Each term in GROUP 1 was cross-tabulated with all terms in GROUP 2. Given the broad scope of the review, we adopted an iterative process and refined the search strategy as we progressed. Key papers were also searched for in databases to identify subject headings or descriptors applied to them, which were then used to further refine the search strategy. The approaches above returned a final search strategy which is explained in Supplementary Material **Table S1**. For all searches, high-income countries and RCT, experimental and quasi-experimental studies, filters were used.

### Eligibility criteria

Overall, only studies (both published and unpublished) from high-income countries, as defined by the World Bank, providing impact evidence of income-support interventions on the outcomes of interest were included in the review.

Macro-level interventions of interest included all strategies aimed to increase income i.e. income support intervention, among which:

⍰ Social protection strategies (based on social assistance and safety nets, such as: conditional or unconditional cash transfers; price subsidies for electricity, public transport or food such as food stamps, vouchers, and coupons);
⍰ Taxation policies and benefits (i.e. fee waivers and exemptions for schooling, tax credits, and utilities);
⍰ Interventions aimed to enhance and promote lone parent employment, namely Welfare to Work (WtW) interventions, if they included cash benefits that could directly affect outcome;
⍰ Minimum wage salary policies.

Interventions addressing differential exposure to risk factors (i.e. housing) and differential vulnerability to risk factors in disadvantaged groups (i.e. support for disabled people in the household) were *not* included. We did *not* include school feeding programs as they were considered as a separate type of intervention, also typically delivered after the age window of interest. We also did not include interventions that directly affected health outcomes (e.g. Medicaid or medical insurance-related interventions) because they could affect directly child health, beyond our conceptual framework pathways.

We considered interventions delivered during children’s first 1000 days of life only: this is a key period for determining lifetime health trajectories, since influences in early-life can cause long-term functional and structural changes.(19)

Outcomes of interest included childhood life-course risk factors and health outcomes concerning:

⍰ Cardiovascular health (e.g. specific diseases or parameters as blood pressure measurements);
⍰ Metabolic conditions (e.g. birth weight, obesity, diabetes mellitus);
⍰ Respiratory diseases: (e.g. Wheezing, Asthma, COPD);
⍰ Mental health: specific diagnoses (e.g. ADHD, ASD, Internalizing/Externalizing behaviour problems) or self-assessment/reporting of mental health status.

Studies including impact on generic, self-reported measures of the overall health status were not included.

Finally, we applied restrictions on study design including only studies that reported impact evidence from Randomised Controlled Trials (RCTs) and Quasi-Experimental design studies. No time or language restriction was applied to papers.

### Studies selection

Two authors independently performed the selection process for each paper identified through database search or snowballing procedures, following the PRISMA flowchart, in order to assess eligibility; the whole procedure is described in **Figure 2**. Disagreements in the inclusion of the paper were resolved by consensus, consulting the other members of the team if appropriate.

**Figure 2.**
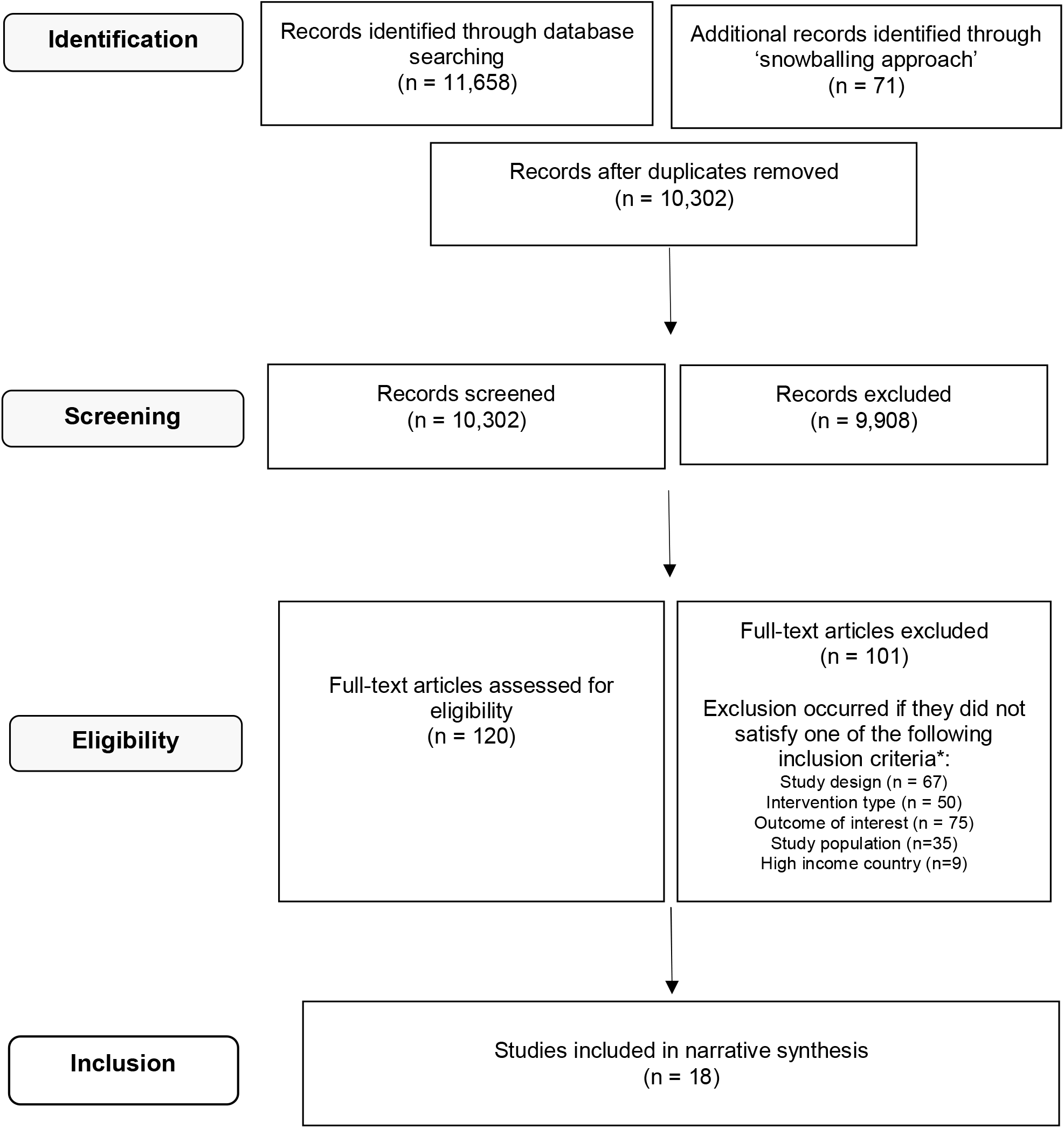
Flow diagram of studies included/excluded by stage. * Some studies were excluded because they did not meet more of the inclusion criteria, therefore they contribute to more of the below categories

### Data extraction appraisal and synthesis

Data extraction forms were created to gather relevant information from the selected papers. Given the anticipated heterogeneity of studies, we did not conduct a meta-analysis. We summarised instead the principal findings of each study and combined them together via a narrative synthesis.

### Risk of bias assessment

Despite the existence of several tools for the critical appraisal of the quality of studies, we chose to use the approach suggested by Waddington et al based on the simple identification of a number of selected biases (whether explicitly stated in the papers or identified by the authors of this report) (18). Due to the complexity and heterogeneity of the studies and their statistical techniques, in particular for quasi-experimental ones, in fact, we did not apply any bias score-based approach to determine the overall risk of bias of the eligible papers,(18) but the identified biases were listed, described, and summarised in a descriptive table (**Table 6**).

### Review protocol registration

The review protocol has been successfully registered within PROSPERO in June 2020 with the registration number CRD42020178543.(20)

## Results

The search strategy returned a total of 11,658 papers. After removing duplicates and titles of no relevance, we obtained 358 papers to submit for abstract screening of which 95 were considered suitable for the eligibility assessment. Furthermore, the snowballing approach returned us 71 papers, of which 25 were considered of potential interest. Thus, in total 120 papers underwent eligibility assessment of which 18 met the review requirements. 102 were excluded because they did not meet one or more inclusion criteria as follows: 67 papers did not satisfy the study design; 50 the intervention; 75 did not include the health outcome of interests; 35 were not suitable due to the study population and 9 did not refer to high income countries **(Figure 2)**. Among the 18 eligible papers, 16 referred to independent studies whereas two were a meta-analysis(21) and a pooled analysis(22) of respectively 12 and 5 RCTs concerning Welfare-to-Work interventions.

### Studies description

**Table 2** provides an overview of the main features of the studies included in this review. Their publication year ranged from 2001 to 2019 and the considered interventions were delivered between 1957 and 2013.

**Table 2.**
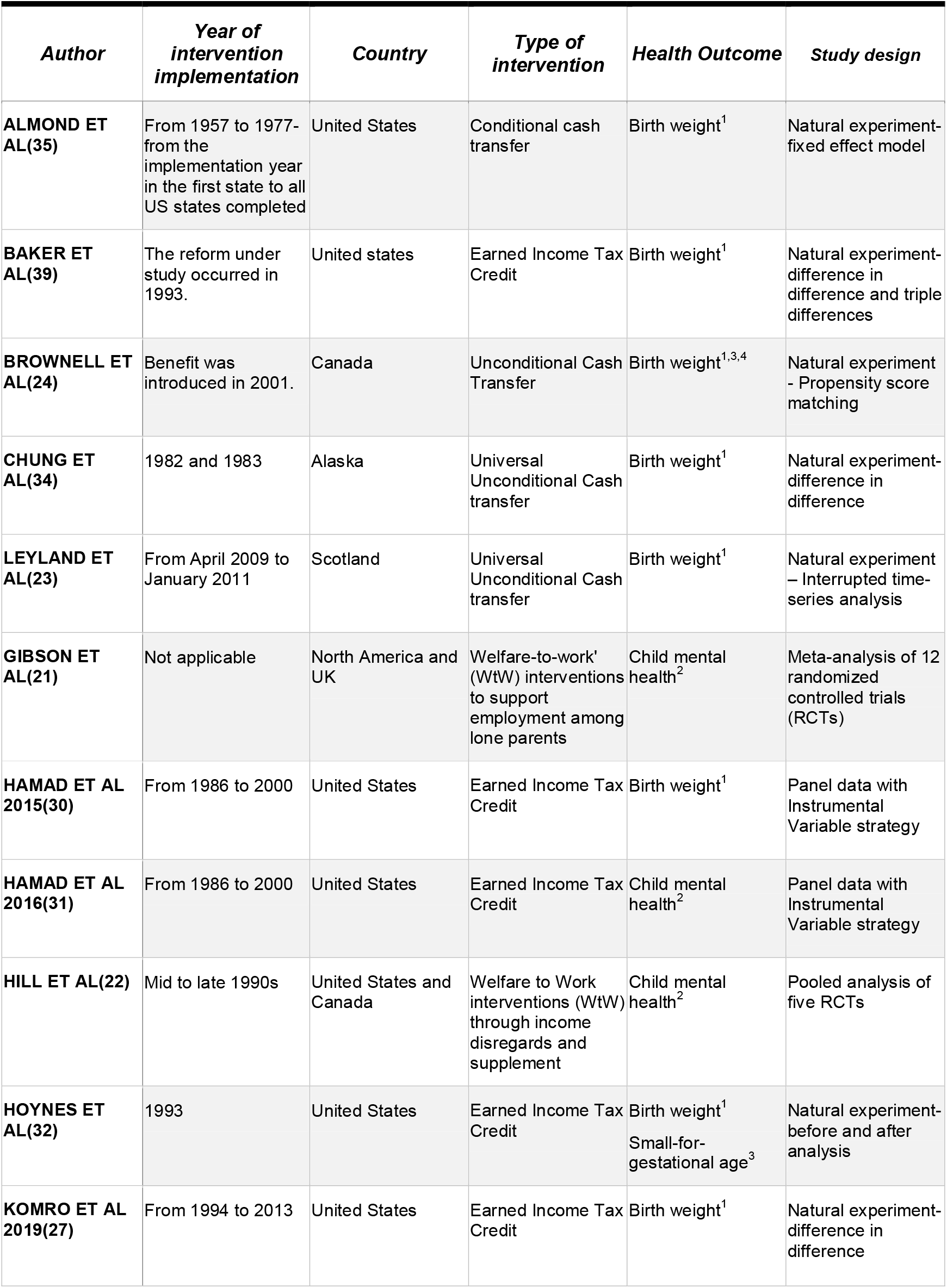
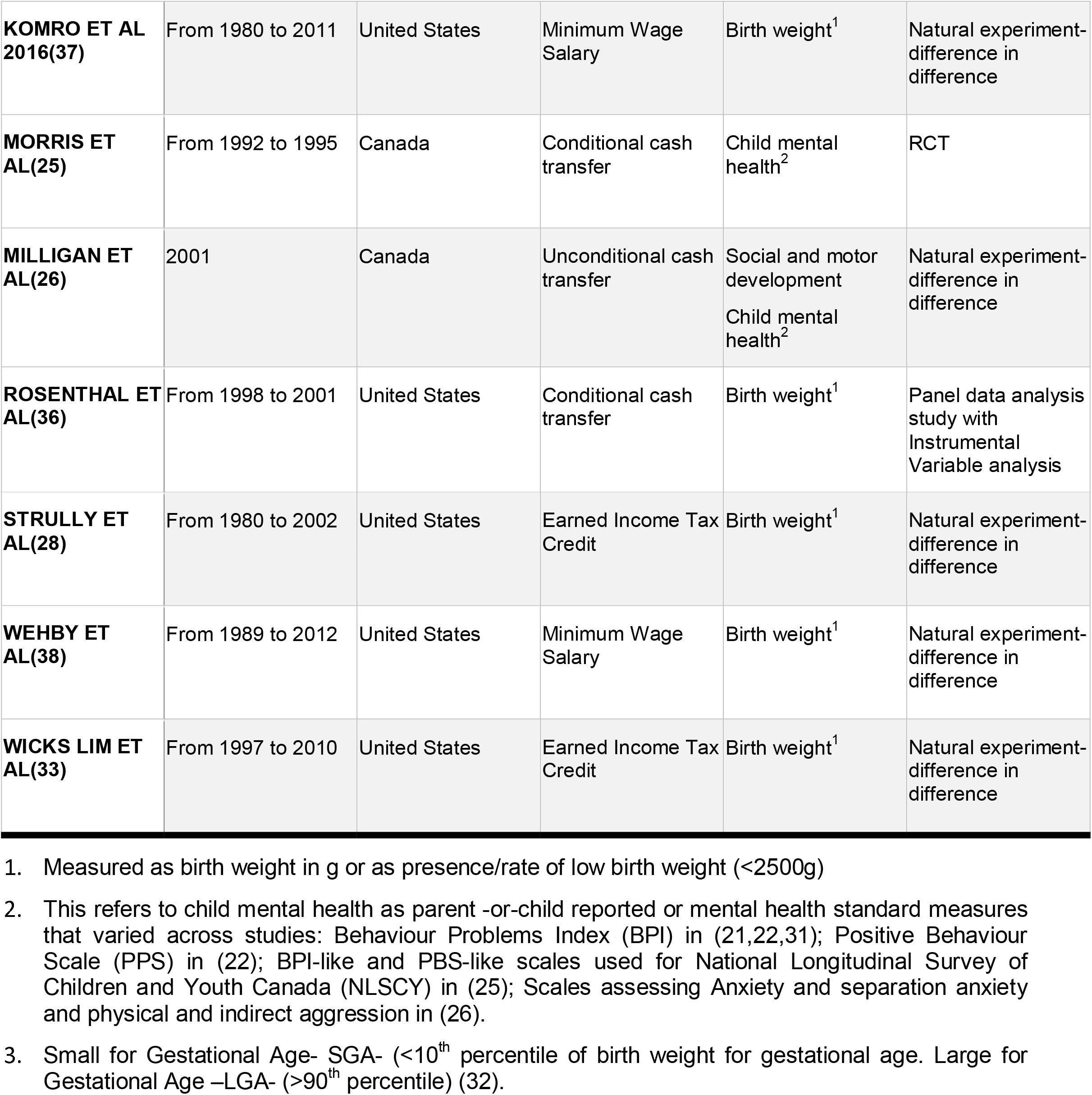
Synopsis of studies included in the review.

| *Author* | *Year of intervention implementation* | *Country* | *Type of intervention* | *Health Outcome* | *Study design* |
| --- | --- | --- | --- | --- | --- |
| **ALMOND ET AL(35)** | From 1957 to 1977- from the implementation year in the first state to all US states completed | United States | Conditional cash transfer | Birth weight<sup>1</sup> | Natural experiment- fixed effect model |
| **BAKER ET AL(39)** | The reform under study occurred in 1993. | United states | Earned Income Tax Credit | Birth weight<sup>1</sup> | Natural experiment- difference in difference and triple differences |
| **BROWNELL ET AL(24)** | Benefit was introduced in 2001. | Canada | Unconditional Cash Transfer | Birth weight<sup>1,3,4</sup> | Natural experiment - Propensity score matching |
| **CHUNG ET AL(34)** | 1982 and 1983 | Alaska | Universal Unconditional Cash transfer | Birth weight<sup>1</sup> | Natural experiment- difference in difference |
| **LEYLAND ET AL(23)** | From April 2009 to January 2011 | Scotland | Universal Unconditional Cash transfer | Birth weight<sup>1</sup> | Natural experiment – Interrupted time-series analysis |
| **GIBSON ET AL(21)** | Not applicable | North America and UK | Welfare-to-work' (WtW) interventions to support employment among lone parents | Child mental health<sup>2</sup> | Meta-analysis of 12 randomized controlled trials (RCTs) |
| **HAMAD ET AL 2015(30)** | From 1986 to 2000 | United States | Earned Income Tax Credit | Birth weight<sup>1</sup> | Panel data with Instrumental Variable strategy |
| **HAMAD ET AL 2016(31)** | From 1986 to 2000 | United States | Earned Income Tax Credit | Child mental health<sup>2</sup> | Panel data with Instrumental Variable strategy |
| **HILL ET AL(22)** | Mid to late 1990s | United States and Canada | Welfare to Work interventions (WtW) through income disregards and supplement | Child mental health<sup>2</sup> | Pooled analysis of five RCTs |
| **HOYNES ET AL(32)** | 1993 | United States | Earned Income Tax Credit | Birth weight<sup>1</sup><br>Small-for-gestational age<sup>3</sup> | Natural experiment- before and after analysis |
| **KOMRO ET AL 2019(27)** | From 1994 to 2013 | United States | Earned Income Tax Credit | Birth weight<sup>1</sup> | Natural experiment- difference in difference |
| **KOMRO ET AL**<br>**2016(37)** | From 1980 to 2011 | United States | Minimum Wage<br>Salary | Birth weight<sup>1</sup> | Natural experiment-<br>difference in<br>difference |
| **MORRIS ET**<br>**AL(25)** | From 1992 to 1995 | Canada | Conditional cash<br>transfer | Child mental<br>health<sup>2</sup> | RCT |
| **MILLIGAN ET**<br>**AL(26)** | 2001 | Canada | Unconditional cash<br>transfer | Social and motor<br>development<br>Child mental<br>health<sup>2</sup> | Natural experiment-<br>difference in<br>difference |
| **ROSENTHAL ET**<br>**AL(36)** | From 1998 to 2001 | United States | Conditional cash<br>transfer | Birth weight<sup>1</sup> | Panel data analysis<br>study with<br>Instrumental<br>Variable analysis |
| **STRULLY ET**<br>**AL(28)** | From 1980 to 2002 | United States | Earned Income Tax<br>Credit | Birth weight<sup>1</sup> | Natural experiment-<br>difference in<br>difference |
| **WEHBY ET**<br>**AL(38)** | From 1989 to 2012 | United States | Minimum Wage<br>Salary | Birth weight<sup>1</sup> | Natural experiment-<br>difference in<br>difference |
| **WICKS LIM ET**<br>**AL(33)** | From 1997 to 2010 | United States | Earned Income Tax<br>Credit | Birth weight<sup>1</sup> | Natural experiment-<br>difference in<br>difference |

1. Measured as birth weight in g or as presence/rate of low birth weight (<2500g) 2. This refers to child mental health as parent -or-child reported or mental health standard measures that varied across studies: Behaviour Problems Index (BPI) in (21,22,31); Positive Behaviour Scale (PPS) in (22); BPI-like and PBS-like scales used for National Longitudinal Survey of Children and Youth Canada (NLSCY) in (25); Scales assessing Anxiety and separation anxiety and physical and indirect aggression in (26). 3. Small for Gestational Age- SGA- (<10<sup>th</sup> percentile of birth weight for gestational age. Large for Gestational Age –LGA- (>90<sup>th</sup> percentile) (32).

With the exception of Leyland et al(23) and Gibson et al,(21) that provided evidence from the United Kingdom, all the other studies were conducted in North America: largely from the United States and in four cases from Canada.(22, 24-28)

The interventions largely focused on a United States poverty alleviation strategy, the Earned Income Tax Credit (seven papers)(27-33) followed by unconditional cash transfer interventions (five papers),(23, 24, 26, 34) conditional cash transfers (three papers),(25, 35, 36) Welfare to Work interventions,(21, 22) and minimum wage salary (two papers).(37, 38)

The meta-analysis and the pooled analysis mentioned before(21, 22) were both summarizing evidence on Welfare-to-Work interventions, defined as all ranges of interventions involving financial incentives and sanctions, training, childcare subsidies and lifetime limits on benefit receipt that are used to support or mandate employment among parents.(21)

**Table 3** provides a detailed description of the interventions included in this review both in terms of benefits provided and recipients (i.e. the ideally target population).

**Table 3.**
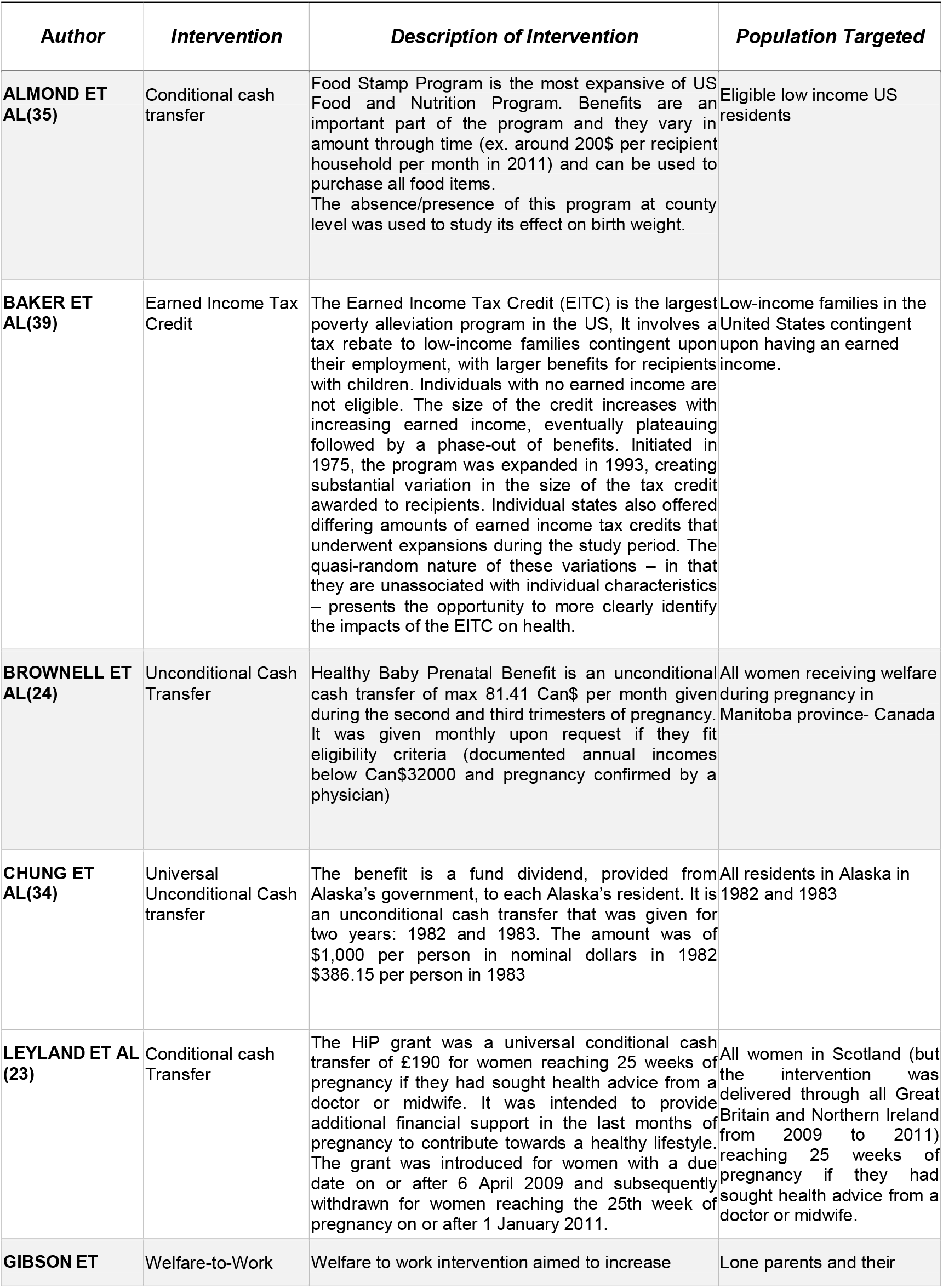
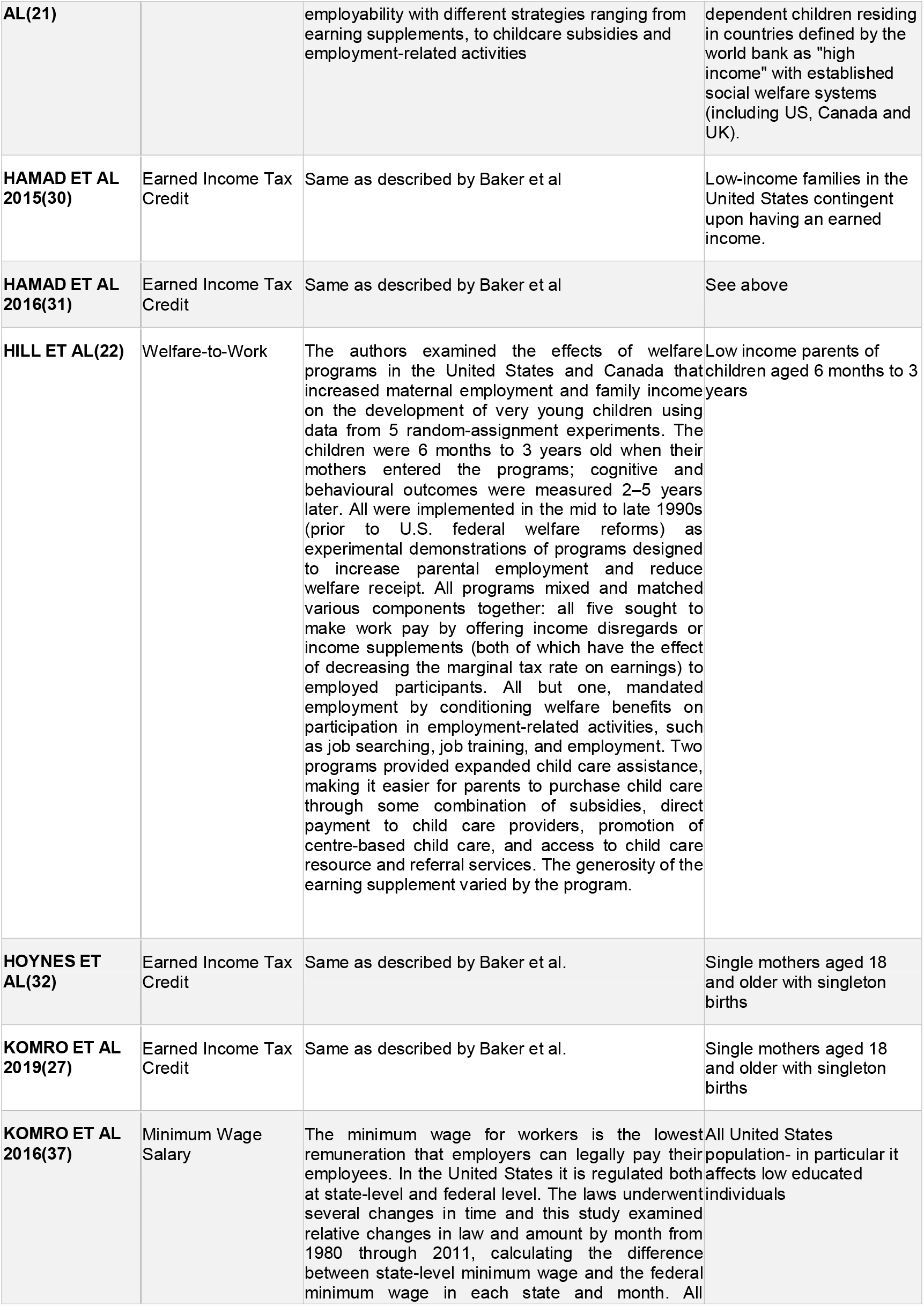
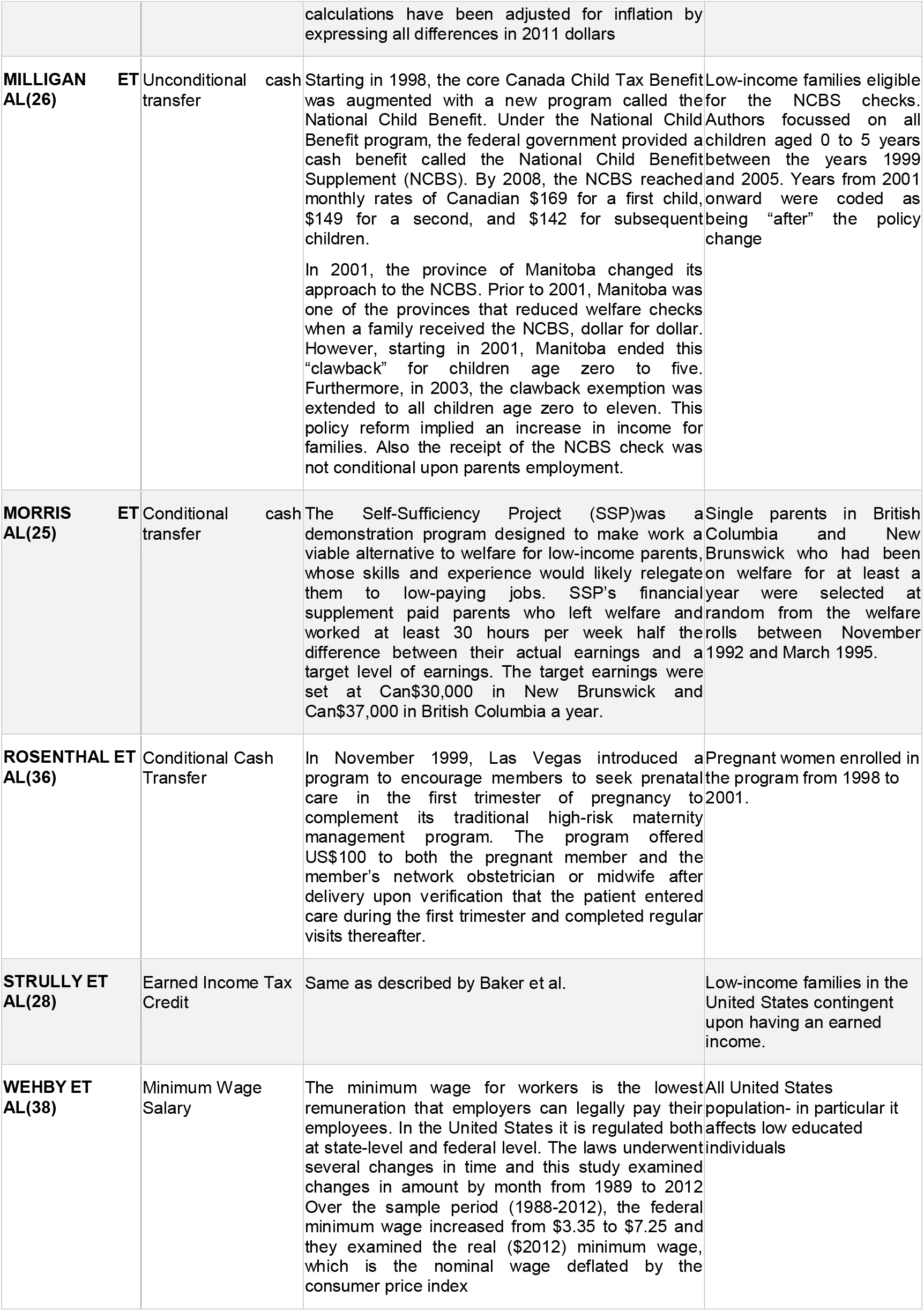
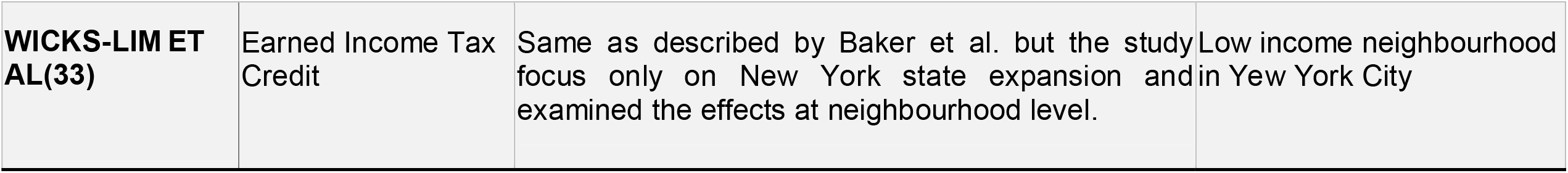
Intervention description by type, theory of change and target population.

In terms of child health outcomes (Table 2), the vast majority of studies focused on birth weight as both its measurement in grams and low birth weight percentage. In two studies,(24, 32) also weight-for-gestational age was examined. In five cases, authors focused on child mental health, quantified either as a measure of mental health on a 1 to 5 scale reported from parents, or through specific instruments, namely the Behavior Problem Index (BPI),(21, 22, 31) the Positive Behaviour Scale, (PBS)(22) some country-specific modified version of them (25) and the anxiety and physical and indirect aggression scale.(26)

Studies have largely relied on quasi-experimental study designs, whereas RCTs have been considered only in the meta and pooled-analyses(21, 22) and in one conditional cash transfer from Canada **(Table 3)**.(25) Quasi-experimental designs adopted a wide range of impact evaluation techniques of different rigour and complexity: before and after analysis,(32) difference in difference,(26-29, 33, 34, 37, 38) instrumental variable analysis,(30, 31) interrupted time series analysis,(23) propensity score matching,(24) and Fixed Effect Model.(35)

### Effect findings

We reported impact findings both qualitatively (**Table 4 and Table 5**) and quantitatively (**Table S2**): if the considered paper provided one main model we referred to it, while if the study adopted multiple models or subgroups, all findings’ directions were reported in the tables.

**Table 4.**
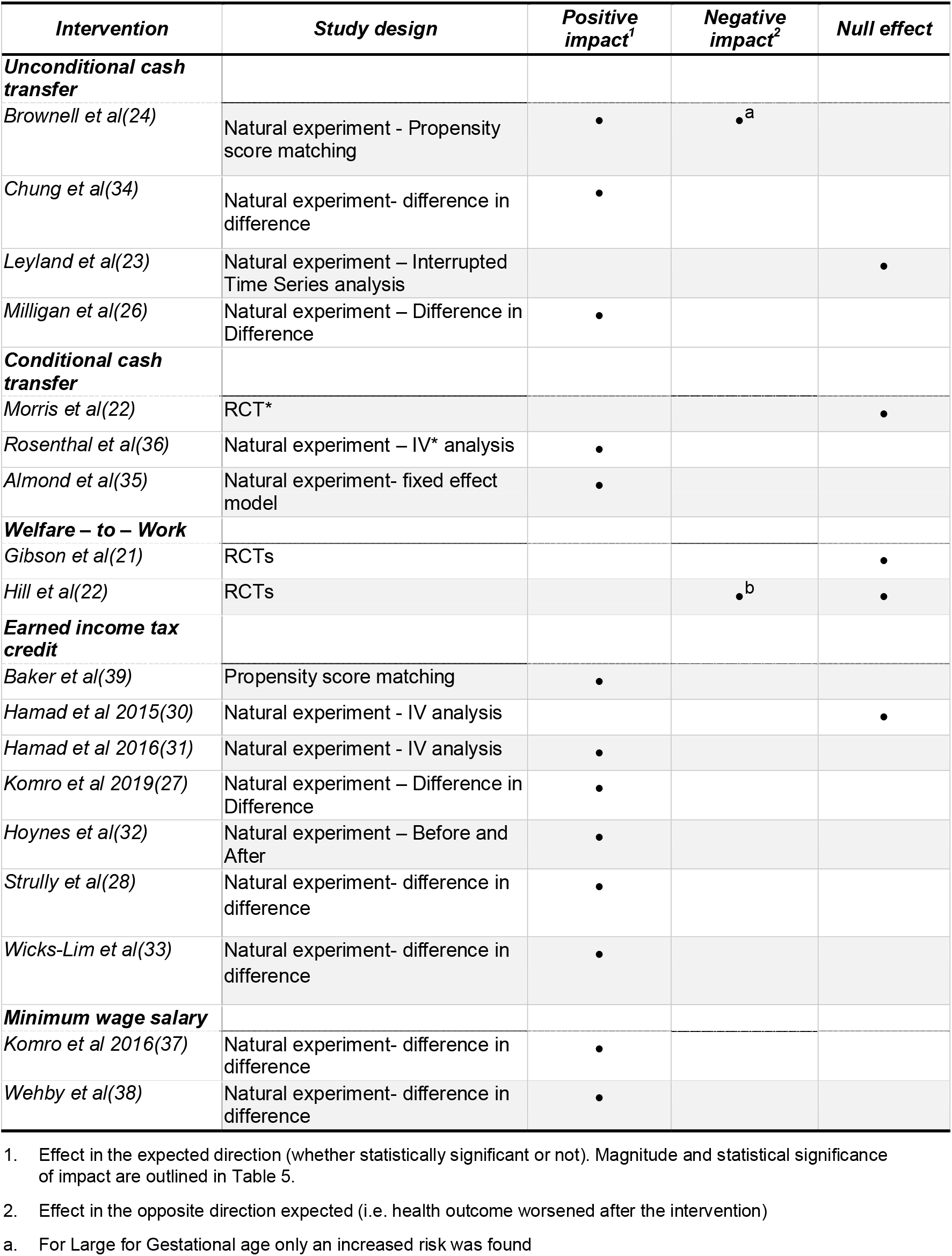
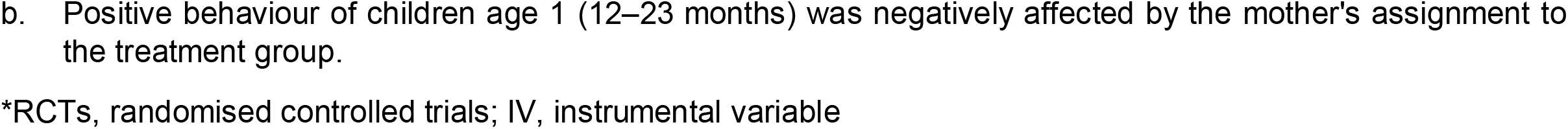
Effect findings by study design and type of intervention.

| *Intervention* | *Study design* | *Positive impact<sup>1</sup>* | *Negative impact<sup>2</sup>* | *Null effect* |
| --- | --- | --- | --- | --- |
| **Unconditional cash transfer** |  |  |  |  |
| *Brownell et al(24)* | Natural experiment - Propensity score matching | • | •<sup>a</sup> |  |
| *Chung et al(34)* | Natural experiment- difference in difference | • |  |  |
| *Leyland et al(23)* | Natural experiment – Interrupted Time Series analysis |  |  | • |
| *Milligan et al(26)* | Natural experiment – Difference in Difference | • |  |  |
| **Conditional cash transfer** |  |  |  |  |
| *Morris et al(22)* | RCT* |  |  | • |
| *Rosenthal et al(36)* | Natural experiment – IV* analysis | • |  |  |
| *Almond et al(35)* | Natural experiment- fixed effect model | • |  |  |
| **Welfare – to – Work** |  |  |  |  |
| *Gibson et al(21)* | RCTs |  |  | • |
| *Hill et al(22)* | RCTs |  | •<sup>b</sup> | • |
| **Earned income tax credit** |  |  |  |  |
| *Baker et al(39)* | Propensity score matching | • |  |  |
| *Hamad et al 2015(30)* | Natural experiment - IV analysis |  |  | • |
| *Hamad et al 2016(31)* | Natural experiment - IV analysis | • |  |  |
| *Komro et al 2019(27)* | Natural experiment – Difference in Difference | • |  |  |
| *Hoynes et al(32)* | Natural experiment – Before and After | • |  |  |
| *Strully et al(28)* | Natural experiment- difference in difference | • |  |  |
| *Wicks-Lim et al(33)* | Natural experiment- difference in difference | • |  |  |
| **Minimum wage salary** |  |  |  |  |
| *Komro et al 2016(37)* | Natural experiment- difference in difference | • |  |  |
| *Wehby et al(38)* | Natural experiment- difference in difference | • |  |  |

1. Effect in the expected direction (whether statistically significant or not). Magnitude and statistical significance of impact are outlined in Table 5.
2. Effect in the opposite direction expected (i.e. health outcome worsened after the intervention)
a. For Large for Gestational age only an increased risk was found
- b. Positive behaviour of children age 1 (12–23 months) was negatively affected by the mother's assignment to the treatment group.
\*RCTs, randomised controlled trials; IV, instrumental variable

**Table 5.**
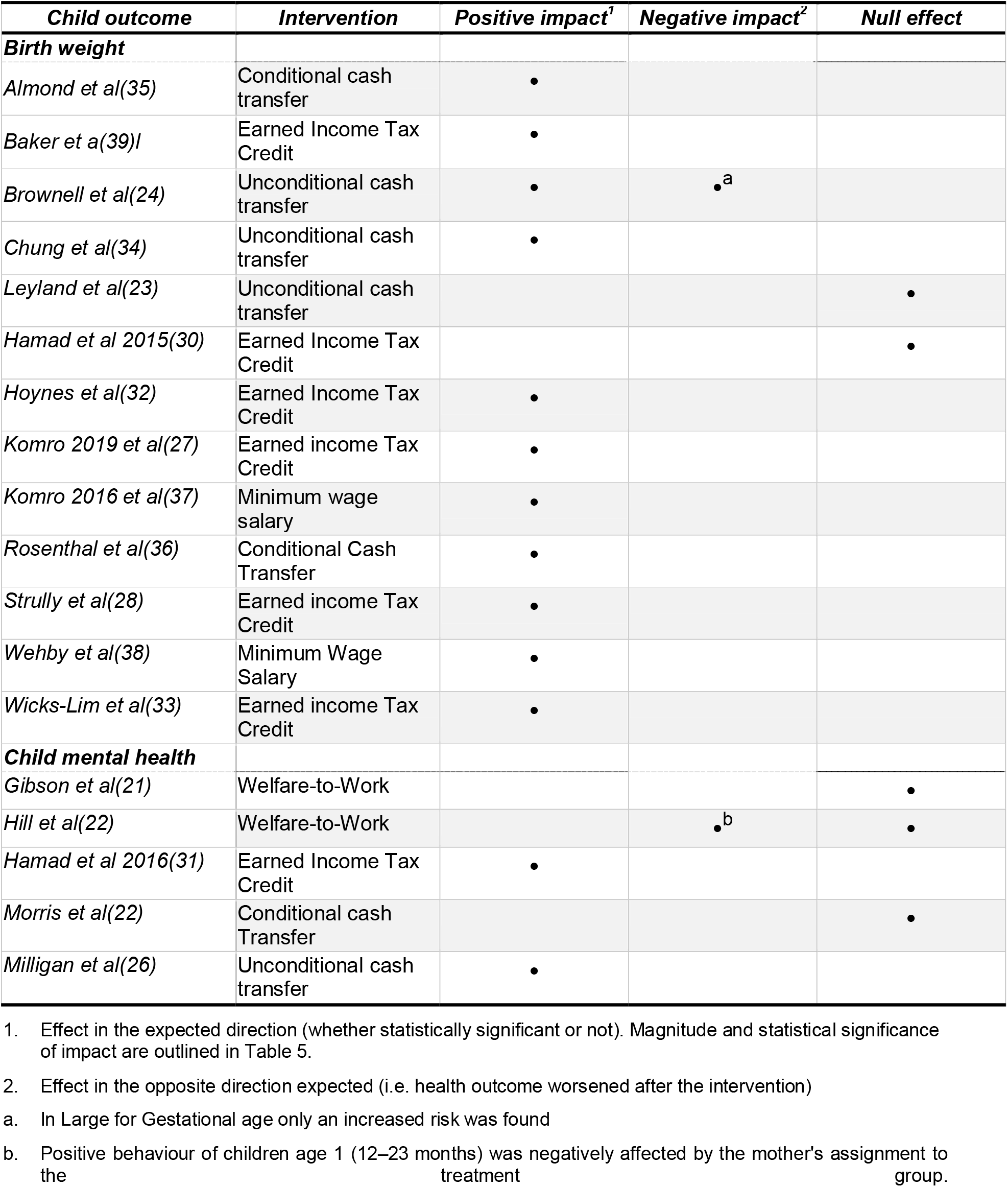
Effect findings by child health outcome.

Findings have been labelled as positive, when demonstrating a positive effect in the expected direction (i.e. health outcome improvement), regardless of issues of statistical significance, negative when showing an impact in opposite direction expected (i.e. health outcome worsening) or null, when no effect or any clear direction was observed.

As shown in **Table 4 and 5**, for almost all the considered interventions results seem to lean toward an overall positive effect on the considered outcome, with some exceptions. Null results were observed in six studies. In two cases(22, 24) authors documented a negative impact of the intervention: in Hill et al,(22) the positive behaviour among children in age group 12-23 months was negatively affected by mothers’ assignment to the treatment group, while in Brownell et al an increased risk of Large for

Gestational Age (LGA) was found for children whose families received an unconditional cash transfer during pregnancy in Canada.(24) In both instances negative results were paralleled with positive or null findings in other subgroup analyses.(22, 24)

#### Findings by intervention type

**Table 4** provides a distribution of the impact findings by type of intervention.

Welfare to Work interventions, overall, had no effect on the given outcome (i.e. mental health). The same applies to the RCT on the Self Sufficiency Project (SSP),(25) a conditional cash transfer aimed to increase employment, which provided a financial supplement to parents who left welfare and worked at least 30 hours per week.

The Earned Income Tax credit, conducted in the United States,(27, 28, 30-33, 39) by contrast seems to produce almost consistently a positive impact on child birth weight, in six of the seven quasi-experimental studies that examined it, except for Hamad 2015 irrespective of the statistical method adopted.(30, 31)

Finally, the remaining unconditional and conditional cash transfer interventions examined showed overall a positive impact on child health, both on reduction of absolute birth or low birth weight prevalence and on children’s mental health scores.

### Findings by study design and health outcomes

Results did not appear to be robust to the study design adopted **(Table 5)**, with all RCTs consistently reporting null or negative effect.(21, 22, 25) Contrarily, all quasi-experimental studies, except for Milligan et al(26) and Hamad et al,(30) found at least one positive effect.

Despite the differences in the number of studies tackling birth weight and mental health (i.e. respectively 13 vs 5), evidence seems to suggest a more consistent response from studies looking at birth weight compared to those looking at mental health (**Table 5**).

Specifically, studies looking at mental health showed a more mixed picture, with positive and null effects almost equally represented among studies: the two RCTs, concerning Welfare to Work intervention and the one on the SSP program, designed to promote work among low income families did not find any effect on mental health. Conversely the other two quasi-experimental studies concerning respectively the EITC and the Canadian Child Tax Benefit, found a positive effect of the interventions on children’s mental health. Evidence on birth weight offered better consistency with findings being more aligned and showing overall a clearer positive trend.

### Magnitude of positive effects

Different study designs and result presentations adopted do not allow us to standardise the magnitude of the observed impact, but all the quantitative findings are reported in Supplementary Material **Table S2**.

Given the high heterogeneity of studies involved and the modest impact size, a comparison of the magnitude of impact across different interventions is of limited meaning and the population health implications remain uncertain. Nonetheless, in two cases,(27, 37) authors attempted to extrapolate the observed effect into actual negative public health outcomes averted. According to authors,(27) the 12% reduction in low birth weight found for EITC translates into 3760 fewer low birth weight babies born from black mothers and 8364 fewer low birth weight babies born from white mothers per year across the United States. Hispanic and non-Hispanic mothers displayed relatively similar effects. For minimum wage salaries instead, if all United States in 2014 had increased their minimum wages by 1 dollar there would likely have been an estimated 2790 fewer low birth weight births for the year.(27)

### Conceptual frameworks

The majority of papers included in this review, except for(24, 33, 37), explicitly mentioned a theory of change or logic model either informing their study hypotheses or guiding the results interpretation. Multiple pathways were speculated through which those interventions, aimed at income support, could affect perinatal health if delivered during pregnancy.

Health-related behaviours were predominantly mentioned, mainly smoking, alcohol, and consumption unhealthy foods that are unevenly distributed across different socioeconomic positions. Those behaviours can directly affect infant health, acting in particular on intra-uterine growth that eventually is a key determinant of birth weight.(35)

In addition, women with lower household income suffer from higher rates of malnutrition, demonstrate heightened psychological stress associated with neuroendocrine dysfunction, which can ultimately influence the likelihood and duration of breastfeeding and hamper access to adequate prenatal care services.(30) Maternal healthcare utilisation behaviours (prenatal care), in particular, was analysed in three studies(32, 34, 36) suggesting some evidence for a mediating role.

Some studies also mentioned the “family process” conceptual model,(25, 27, 28, 30-32) which postulates that the extra income provided by child benefits may improve long-run outcomes, not only through direct investments but by improving also the emotional environment in which the children grow.(22) Specifically, maternal depression and parental warmth were both identified as potential mediators of welfare programs’ impact in most studies.(21, 22, 25, 27, 28, 30, 32, 38) In all these studies, income and employment were hypothesised to affect parental mental health which in turn affected child physical and mental health.

### Quality assessment of the studies

In all the assessed studies, at least one bias type was detected. According to Waddington et al, we reported on **Table 6** the identified biases if declared in the paper or detected by one of the authors of this review. The overall study quality was moderate-high, and they often dealt with complex analyses and multiple statistical tools. However, due to the complexity of evaluating such interventions, the majority of the studies were at risk of exposure misclassification, either differential or non-differential. Furthermore, most of the studies were affected by incomplete reporting because of their lack in sharing either some results or, in most cases, relevant information for their interpretation, such as how they dealt with missing data.

**Table 6.** Biases identified in the studies included in the review.

|  | Information bias | Misclassification | Omitted variable bias | Reporting bias | Selection bias | Other |
| --- | --- | --- | --- | --- | --- | --- |
| *Description* | Differences in the collection, recall, recording or handling of information used in a study | Incorrect classification of participants into categories | The statistical model leaves out one or more relevant variables | Selective disclosure or withholding of information by parties involved in the design, conduct, analysis, or dissemination | Individuals or groups in a study differ systematically from the population of interest |  |
| Almond et al(35) |  | • |  | • |  |  |
| Baker et al(39) |  | • | • |  |  |  |
| Brownell et al(24) |  | • |  |  | • a | • a |
| Chung et al(34) | • | • |  |  |  |  |
| Gibson et al(21) |  |  |  |  | • | • b |
| Hamad et al 2015(30) | • | • |  |  | • | • c |
| Hamad et al 2016(31) | • | • |  |  |  |  |
| Hill et al(22) |  |  |  | • |  |  |
| Hoynes et al(32) |  | • | • | • |  |  |
| Komro et al 2016(37) |  | • |  | • |  |  |
| Komro et al 2019(27) |  |  |  |  | • | • d |
| Leyland et al(23) | • |  |  | • |  |  |
| Milligan et al(26) |  |  |  | • | • |  |
| Morris et al(22) |  |  |  | • |  |  |
| Rosenthal et al(36) |  |  | • | • |  |  |
| Strully et al(28) |  | • |  |  |  | • e |
| Wehby et al(38) |  | • |  | • |  |  |
| Wicks-Lim et al(33) | • | • |  |  |  | • f |

a. They could not ascertain how much unmeasured confounding influenced results, causing endogeneity bias. Furthermore, they limited the evaluation to women receiving welfare rather than examining all low-income women receiving the income supplement during pregnancy, thus limiting the generalizability of results.
b. Due to the heterogeneity of the studies their comparison could be unspecific and
c. Recall bias may be present from women interviewed not in the year of childbirth
d. Observed result of LBW could be an underestimation of the differential effect by race because black women had a double rate of stillbirths
e. Mother's life environment and attitude or unmeasured genetic variation could influence results
f. Some factors that could cause spurious correlation such as neighbourhood gentrification, were impossible to control for.

## Discussion

This review aimed to quantify the impact of income-support strategies on life-course risk factors and health outcomes. This review adds to the existing literature by providing insights on the impact of specific types of macro-economic interventions on a specific window period (i.e. first 1,000 days of life) and a selected list of specific life-course risk factors and health outcomes. Consistent with similar evidence synthesis efforts,(21, 40) in this review we could not conclusively demonstrate an effect of income-support-strategies on all the selected child outcomes. Overall, evidence available suggest that income support strategies have a positive, albeit small, effect on birth weight and limited impact on mental health indicators. No other health outcome of interest was investigated in the studies included in this review, so no inference can be made on cardiometabolic and respiratory health outcomes.

One could argue that despite the small observed effect, the proportion of people exposed to these policies is quite large which could result overall into a considerable effect in public health terms. Nonetheless, only two studies in this review have tried to extrapolate this effect to the population level. (27, 32)

The general conclusion of this review seems to be robust to the type of intervention and child outcome under observation, but not to the study design, with quasi-experimental studies consistently more likely to detect a positive impact than randomised controlled trials.

There are possible, not mutually exclusive, explanations for the results of our review, including the fact that despite the extensive screening of different, multidisciplinary literature browsers, the search strategy returned a relatively small number of eligible studies. This is consistent with the conclusion of similar previous reviews(21, 40)claiming that evidence are inconclusive also because very few experimental or quasi-experimental studies have been undertaken to explore the impact of complex, macro-level socioeconomic interventions on child health and - even less - on specific, well measured child health outcomes. We restricted the review to well-defined life-course risk factors and health outcomes, thus excluding studies assessing the effects on generally self-reported health or well-being, namely those subjective outcomes that are strongly affected by both non-differential and differential misclassification. This selection has however further limited the literature to draw upon. The limited effect on mental health outcomes could be also due to the poor standardisation in the definition and measurement of these outcomes.

If we exclude studies that have used a randomised control study designs,(21, 22, 25) results appear less mixed and more convincingly leaning towards an overall positive effect: as argued by other authors,(41, 42) thus, quasi-experimental studies seem to be more suitable to the evaluation of complex interventions with weak effects in a large group of the population. Given the limited number of RCTs included in our review, this remains speculative, but there is clearly the need to understand how methodological aspects influence our understanding of the health impact of these policies.

It is worth noting that with few exceptions,(23, 36) most of the interventions included in the review were not originally designed and implemented to evaluate nor achieve a health effect. This implies that some of the potential impact of these programs could have been missed purely for design/implementation reasons. On the other hand, for those interventions that had a quantifiable effect (e.g. the Earned Income Tax credit studies), one could argue how bigger this effect could have been if these programs were designed with the precise intent of improving people health other than just socioeconomic measures.

The relative modest effect observed in the studies included in this review could be attributed to the size of the benefits provided: this may explain for example why the impact of the Earned Income Tax credit (where the size of cash received can be relatively high (31, 32)) seems almost consistently positive. By contrast, the two studies on conditional cash transfers included in this review, involving a fairly small overall cash transfer provided to beneficiary women of 190GBP or 100 USD, found no evidence of an effect.(23, 36) These observations are consistent with what reported in other reviews similar to this one. For example, Lucas at al(40) concluded that the monetary value of many interventions was low, as in most studies included in their review the total increase in income to intervention families was less than US$50 per month despite the fact that many parents were compelled to work full-time.(22) Authors questioned whether the level of income increase was sufficient to affect living conditions and – we would add – it was big enough to ensure this effect translated into a health effect.(40) Similarly the impact of Welfare-to-work interventions on health were unlikely to have a tangible impact and this largely because of the small effect on the economic outcomes (i.e. income).(21) Authors observed that even where employment and income were increased by the intervention for lone parents enrolled in these programs, poverty was still high for most of them in many of the studies. Perhaps because of this, depression also remained very high for lone parents whether they were enrolled in these programs or not.(21)

Most of the interventions included in this review focus on indicators of socioeconomic position or - broadly speaking – econometric concepts of disadvantage. While the association between these constructs and child health is widely acknowledged, this relationship is likely to be complex and mediated by a number of underlying known and unknown pathways. Importantly, if the effect on income does not translate into a tangible effect on these mediators then the expected impact on child health may not materialise as expected. As in Gibson et al,(21) the role of parental depression and in particular maternal mental health has been speculated to be an important mediator. Other studies(26, 32) suggest a ‘family process’ mediation pathway according to which the extra income provided by the child benefits may improve in the long-run outcomes not only through direct financial investment, but also by improving the emotional environment in which children grow up. Another important mediator is whether the increase of income happens via the mother’s employment:(22, 32) some authors speculated that some policies that incentivise maternal employment may involuntarily increase maternal stress and add extra pressure on mothers which offsets the benefit of a better income on children. Similarly, Morris et al(25) argue that a proper evaluation of the impact of better income and parental employment on child health should account for the moderating role of the developmental period of the child. According to these authors,(25) the effect of income and employment on children aged 1 or less may be counterbalanced, if not reverted, via prolonged periods of time of maternal absence that ultimately leads to increased instability of care and reduced parental warmth.(25)

Our review presents with a number of limitations. Despite our comprehensive search of the literature, the evidence we gathered provides at most a partial representation of existing macroeconomic policies. This is mostly due to the limited number of health outcomes under investigation and to the heavy predominance of studies from North America, largely focussing on Earned Income Tax Credit in the United States. This unbalance is probably largely due to the fact that Earned Income Tax Credit is the most important poverty-alleviation strategy in the United States and it is particularly suitable to quasi-experimental impact evaluations because of variation in the distribution of benefits and changes in welfare policy. While our findings are still relevant, their external validity to countries beyond the United States, to different types of interventions, and other health outcomes remain limited.

Not only we found evidence just on birth weight and mental health outcomes, but birth weight outcomes were consistently evaluated though quasi-experimental design studies, whereas half of the studies concerning mental health were RCTs. Thus, we cannot conclude whether the differential impact observed for birth weight and mental health outcomes reflect some genuine intervention-outcome relation or are simply affected by the study design.

In several studies, authors documented a differential impact by race and education level. Sometimes the evaluation of impact by race gave conflicting results.(27, 32, 43) Clearly there is the need to understand better this complex interaction effect.

### Implications for future research

This review provides a useful contribution to the literature on the health effects of social policies. Through the extensive review of the evidence, this research allowed to speculate about possible mechanisms through which these policies may play an effect and why they seem to fail in other circumstances. Finally, through the identification of persisting knowledge gaps, it allowed to draw a research agenda for the future:

First, there is clearly a scope to invest more in the evaluation of the child health impact of macro-level socioeconomic interventions by financing more impact evaluations and by advocating for a better design and implementation of these policies to allow their proper health impact assessment.

Second, the association between income and child health is amply demonstrated. If interventions aiming at improving income do not obtain a commensurate effect on child health outcomes, there is clearly something not working either in the type of intervention provided or in the way we measure this effect. RCTs are considered to be often unfeasible and unethical and unable to capture the complexity of social ‘experiments’.(42). On the other hand, quasi-experimental studies are often imperfect tools that only allow for comparisons between sub-optimal groups.(44) Given the above, there is a mandate to investigate the role of alternative methodologies including observational studies as well as mathematical modelling (i.e. microsimulations) in filling the numerous knowledge gaps still surrounding the impact of socioeconomic interventions on child health.

Thirdly, the question of ‘what works?’ should be more correctly replaced by ‘what works for whom and why?’. There is an urgent need to unpack the effect of these interventions to understand better the reasons for their failure and success. This could be achieved through the design of impact evaluations adopting mixed-methods approach and/or requiring the collection of data to perform rigorous moderation and mediation analyses to explore whether some sub-groups may most benefit from the intervention and through what underlying pathway. Alternatively, and perhaps more conveniently, one could complement reviews like this one, with a “realist” approach, that is a type of literature review in which evidence are mapped against a pre-defined conceptual framework to validate or reject the existence of the speculated underlying pathways linking the interventions with the outcomes of interest.(45) This lens could be applied to the subject of this review and provide important additional explanations on the likely impact of these interventions on child health.

Finally, there is scope to expand this literature review by adding evidence on the long-term impact of these interventions. To the best our knowledge, only few studies have explored the long-term health impact of income support strategies. Studies available(46-48) show consistently a long-term positive impact of the interventions of interest on all health and financial outcomes investigated. Nonetheless, due to the paucity of data, conclusions have to be drawn cautiously. It is also worth exploring the extent to which the way vaster literature from low and middle income can contribute to the understanding of the potential public health impact of income support strategies in high income countries. In other words, there may be merits in creating more connections between low/middle income and high income countries on socioeconomic interventions and explore how lessons can be extrapolated to both environments.(49)

## Conclusions

On the basis of this review we have not been able to establish conclusively whether income support policies delivered in the first 1,000 days of life are able to improve important life-course risk factors and child health outcomes. If we concentrate on birth weight and quasi-experimental studies only, evidence suggest a modest positive effect of these policies. However, the breath and scope of the literature needs to be enriched with additional diversified evidence (in terms of health outcome, country and intervention of interest, and other relevant contextual factors) before a definitive conclusion can be reached and the public health potential of these policies is fully understood. The association between lower income and poorer outcome across all dimensions of child health is strong and consistent across countries and time: the fact that a relatively small number of interventions show a small or null effect should be considered as a “research call” to undertake more and better impact evaluations of these policies, able not only to quantify their effect, but also to provide evidence on what works best, for whom, at what development stage and - most importantly – why.

## Data Availability

All data produced in the present work are contained in the manuscript.

## Author contribution

*<u>DB, LR and CP conceptualised the review objectives and design. DB formulated the search strategy and together with SM performed the papers selection and data extraction and curation. DB and SM completed the first draft and the editing. MR and SL provided supervision and validation of the main results and conclusions of the review. All authors contributed to the interpretation of the review findings and formulation of research and policy recommendations</u>*.

## Declaration of interest

The authors declare that they have no affiliations with or involvement in any organization or entity with any financial interest or non-financial interest (such as personal or professional relationships, affiliations, knowledge or beliefs) in the subject matter or materials discussed in this manuscript.

## Founding source

LIFECYCLE: The LIFE-CYCLE project has received funding from the European Union’s Horizon 2020 research and innovation programme under grant agreement No 733206. This publication reflects only the author’s views and the European Commission is not liable for any use that may be made of the information contained therein.

## Notes

### Competing Interest Statement

The authors have declared no competing interest.

### Funding Statement

The LIFECYCLE project has received funding from the European Union Horizon 2020 research and innovation programme under grant agreement No 733206. This publication reflects only the authors views and the European Commission is not liable for any use that may be made of the information contained therein.

## References

1. UNICEF INNOCENTI. Building the Future: Children and the Sustainable Development Goals in Rich Countries. UNICEF Office of Research - Innocenti; 2017.

2. Taylor-Robinson D, Lai ETC, Wickham S, Rose T, Norman P, Bambra C, et al. Assessing the impact of rising child poverty on the unprecedented rise in infant mortality in England, 2000-2017: time trend analysis. BMJ Open. 2019;9(10):e029424.

3. Narayan A, Cojocaru A, Agrawal S, Bundervoet T, Davalos ME, Garcia N, et al. COVID-19 and Economic Inequality : Short-Term Impacts with Long-Term Consequences. World Bank, Washington D.C. ; 2022.

4. Baird J, Jacob C, Barker M, Fall CH, Hanson M, Harvey NC, et al. Developmental Origins of Health and Disease: A Lifecourse Approach to the Prevention of Non-Communicable Diseases. Healthcare (Basel). 2017;5(1).

5. Pearce A, Dundas R, Whitehead M, Taylor-Robinson D. Pathways to inequalities in child health. Arch Dis Child. 2019;104(10):998–1003.

6. Wickham S, Anwar E, Barr B, Law C, Taylor-Robinson D. Poverty and child health in the UK: using evidence for action. Arch Dis Child. 2016;101(8):759–66.

7. Spencer N, Raman S, O’Hare B, Tamburlini G. Addressing inequities in child health and development: towards social justice. BMJ Paediatr Open. 2019;3(1):e000503.

8. Tomlinson M, Darmstadt GL, Yousafzai AK, Daelmans B, Britto P, Gordon SL, et al. Global research priorities to accelerate programming to improve early childhood development in the sustainable development era: a CHNRI exercise. J Glob Health. 2019;9(3):020703.

9. Jaddoe VWV, Felix JF, Andersen AN, Charles MA, Chatzi L, Corpeleijn E, et al. The LifeCycle Project-EU Child Cohort Network: a federated analysis infrastructure and harmonized data of more than 250,000 children and parents. Eur J Epidemiol. 2020;35(7):709–24.

10. Sow M, De Spiegelaere M, Raynault MF. Evaluating the effect of income support policies on social health inequalities (SHIs) at birth in Montreal and Brussels using a contextualised comparative approach and model family method: a study protocol. BMJ Open. 2018;8(9):e024015.

11. Xiamei Guo XFNS. Housing and Support Services with Homeless Mothers: Benefits to the Mother and Her Children. Author manuscript; available in PMC 2017 Jan 1Published in final edited form as: Community Ment Health J. 2016;52(1):73–83.

12. Yeung WJ, Linver MR, Brooks-Gunn J. How money matters for young children’s development: parental investment and family processes. Child Dev. 2002;73(6):1861–79.

13. Del Boca D, Flinn C, Wiswall M. Household Choices and Child Development. The Review of Economic Studies. 2013;81(1):137–85.

14. Mistry RS, Biesanz JC, Taylor LC, Burchinal M, Cox MJ. Family Income and Its Relation to Preschool Children’s Adjustment for Families in the NICHD Study of Early Child Care. Developmental Psychology. 2004;40(5):727–45.

15. Courtin E, Kim S, Song S, Yu W, Muenning P. Can Social Policies Improve Health? A Systematic Review and Meta-Analysis of 38 Randomized Trials. The Milbank Quarterly. 2020;98(2):297–371.

16. Cooper K, Stewart K. Does Household Income Affect children’s Outcomes? A Systematic Review of the Evidence. Child Indicators Research. 2021;14(3):981–1005.

17. Simpson J, Albani V, Bell Z, Bambra C, Brown H. Effects of social security policy reforms on mental health and inequalities: A systematic review of observational studies in high-income countries. Soc Sci Med. 2021;272:113717.

18. Waddington H, White H, Snilstveit B, Garcia Hombrados J, Vojtkova M, Davies P, et al. How to do a good systematic review of effects in international development: a tool kit. Journal of Development Effectivess. 2012;4(3):359–87.

19. Gluckman PD, Hanson MA, Cooper C, Thornburg KL. Effect of in utero and early-life conditions on adult health and disease. N Engl J Med. 2008;359(1):61–73.

20. Booth A, Clarke M, Dooley G, Ghersi D, Moher D, Petticrew M, et al. The nuts and bolts of PROSPERO: an international prospective register of systematic reviews. Syst Rev. 2012;1:2.

21. Gibson M, Thompson H, Banas K, Lutje V, McKee MJ, Martin SP, et al. Welfare-to-work interventions and their effects on the mental and physical health of lone parents and their children. Cochrane Database Syst Rev. 2018;2018(2):009820.

22. Hill HD, Morris P. Welfare policies and very young children: experimental data on stage-environment fit. Dev Psychol. 2008;44(6):1557–71.

23. Leyland AH, Ouédraogo S, Nam J, Bond L, Briggs AH, Gray R, et al. Evaluation of Health in Pregnancy grants in Scotland: a natural experiment using routine data. Public Health Research. 2017;5(6).

24. Brownell MD, Chartier MJ, Nickel NC, Chateau D, Martens PJ, Sarkar J, et al. Unconditional Prenatal Income Supplement and Birth Outcomes. Pediatrics. 2016;137(6).

25. Morris CPM. Findings from the Self-Sufficiency Project: Effects on children and adolescents of a program that increased employment and income. Journal of Applied Developmental Psychology. 2003;24(2):201–39.

26. Milligan K, Stabile M. Child benefits, maternal employment, and children’s health: Evidence from Canadian child benefit expansions. American Economic Review. 2009;99(2):128–32.

27. Komro KA, Wagenaar AC, Markowitz S, Livingston MD. Effects of State-Level Earned Income Tax Credit Laws on Birth Outcomes by Race and Ethnicity. Health Equity. 2019;3(1):61–7.

28. Strully KW, Rehkopf DH, Xuan Z. Effects of Prenatal Poverty on Infant Health: State Earned Income Tax Credits and Birth Weight. Am Sociol Rev. 2010;75(4):534–62.

29. Baker M, Milligan K. Maternity leave and children’s cognitive and behavioral development. Journal of Population Economics. 2015;28(2):373–91.

30. Hamad R, Rehkopf DH. Poverty, Pregnancy, and Birth Outcomes: A Study of the Earned Income Tax Credit. Author manuscript; available in PMC 2015 Sep 1Published in final edited form as: Paediatr Perinat Epidemiol. 2015;29(5):444–52.

31. Hamad R, Rehkopf DH. Poverty and Child Development: A Longitudinal Study of the Impact of the Earned Income Tax Credit. Am J Epidemiol. 2016;183(9):775–84.

32. Hoynes H, Miller D, Simon D. Income, the earned income tax credit, and infant health. American Economic Journal: economic policy. 2015;7(1):172–211.

33. Wicks-Lim J, Arno PS. Improving population health by reducing poverty: New York’s Earned Income Tax Credit. SSM Popul Health. 2017;3:373–81.

34. Chung W, Ha H, Kim B. Money transfer and birth weight: evidence from the Alaska permanent fund dividend.. Economic Inquiry. 2016;54(1):576–90.

35. Almond D, Hoynes HW, Schanzenbach DW. Inside the war on poverty: the impact of food stamps on birth outcomes.. The Review of Economics and Statistics. 2011;93(2):387–403.

36. Rosenthal MB, Milstein A, Li Z, Robertson AD. Impact of Financial Incentives for Prenatal Care on Birth Outcomes and Spending. Health Serv Res. 2009;44(5 Pt 1):1465–79.

37. Komro KA, Livingston MD, Markowitz S, Wagenaar AC. The Effect of an Increased Minimum Wage on Infant Mortality and Birth Weight. Am J Public Health. 2016;106(8):1514–6.

38. Wehby GL, Kaestner R, Lyu W, Dave DM. Effects of the Minimum Wage on Child Health. American Journal of Health Economics. 2022;8(3):412–48.

39. Baker KB, editor Do Cash Transfer Programs Improve Infant Health: Evidence from the 1993 Expansion of the Earned Income Tax Credit1993.

40. Lucas PJ, McIntosh K, Petticrew M, Roberts H, Shiell A. Financial benefits for child health and well-being in low income or socially disadvantaged families in developed world countries. Cochrane Database Syst Rev. 2008(2):CD006358.

41. Bonell C, Hargreaves J, Strange V, Pronyk P, Porter J. Should structural interventions be evaluated using RCTs? The case of HIV prevention. Soc Sci Med. 2006;63(5):1135–42.

42. Petticrew M, Chalabi Z, Jones DR. To RCT or not to RCT: deciding when ‘more evidence is needed’ for public health policy and practice. J Epidemiol Community Health. 2012;66(5):391–6.

43. L Dubay GMKTJRK. Changes in prenatal care timing and low birth weight by race and socioeconomic status: implications for the Medicaid expansions for pregnant women. Health Serv Res. 2001;36(2):373–98.

44. Craig P, Cooper C, Gunnell D, Haw S, Lawson K, Macintyre S, et al. Using natural experiments to evaluate population health interventions: new Medical Research Council guidance. J Epidemiol Community Health. 2012;66(12):1182–6.

45. Pawson R, Greenhalgh T, Harvey G, Walshe K. Realist review--a new method of systematic review designed for complex policy interventions. J Health Serv Res Policy. 2005;10 Suppl 1:21–34.

46. Aizer A, Eli S, Ferrie J, Lleras-Muney A. The Long-Run Impact of Cash Transfers to Poor Families. Am Econ Rev. 2016;106(4):935–71.

47. Hoynes H, Schanzenbach DW, Almond D. Long-Run Impacts of Childhood Access to the Safety Net. American Economic Review. 2016;106(4):903–34.

48. Bailey MJ, Hoynes HW, Rossin-Slater M, Walker R. Is the Social Safety Net a Long-Term Investment? Large-Scale Evidence from the Food Stamps Program. Working paper: 26942. National Bureau of Economic Research; 2020.

49. Siddiqi SPARAM. Do cash transfer programmes yield better health in the first year of life? A systematic review linking low-income/middle-income and high-income contexts. Archives of Disease in Childhood. 2018;103(10):920–6.

