## Supplementary material for "The impact of income-support interventions on life course risk factors and health outcomes during childhood: a systematic review in high income countries": Table S1, Table S2: SUPPLEMENTAL MATERIAL_PREPRINT SUBMISSION.htm

**Table S1 � Final search strategy adopted in this
review**

|  |  |  |
| --- | --- | --- |
| **Search** |  | **Terms** |
| **#1** | **Interventions** | social programmes OR social protection OR government programmes OR government transfers OR cash OR food OR in-kind transfers |
| **#2** |  | child grants OR child benefits OR child allowances |
| **#3** |  | Tax benefits OR Tax exemptions OR Tax credit OR Fiscal |
| **#4** |  | Work-based programs |
| **#5** |  | OR #1 to #4 |
| **#6** | **Health outcome** | Health OR Resp\* Health OR Wheezing OR Resp\* Infections OR Mental OR Behav\* OR Psycho\* OR Cardio\* OR BMI OR Weight OR Obesity OR Blood Pressure OR Lipid\* OR Glycem\* |
| **#7** | **Population** | Prenatal OR Antenatal OR Perinatal OR pregnancy OR pregnant OR mother\* OR parent\* |
| **#8** |  | Postnatal OR Perinatal OR Postpartum |
| **#9** |  | �Infant" OR "Newborn" OR "Child, preschool" OR "childhood" OR "children" |
| **#10** |  | OR #7 to #9 |
| **#11** |  | #5 AND #6 AND #10 |

  

**Table
S2 � Magnitude of positive effect and results interpretation^a^**

|  |  |  |  |  |  |
| --- | --- | --- | --- | --- | --- |
| **Child outcome** | **Intervention** | **Outcome description** | **Measure of impact** | **Effect estimate** | **Results interpretation** |
| ***Birth weight*** |  | | | | |
| **Almond et al****(35)** | Conditional cash transfer | Birth weight in grams   Low Birth Weight percentage (LBW%) | Regression coefficient   Percentage point difference | **Birth weight**  (1) Whites^2^: 2.039 p<0.10� Blacks: 3-454  (2) Whites: 2.635 p<0.05 Blacks: 4.120  (3) Whites: 2.089 p<0.10 Blacks: 5.466  (4) Whites: 2.175 p<0.05 Blacks: 1.665  **LBW%**  (1) Whites: -0.0006 p<0.10� Blacks: -0.0015  (2) Whites: -0.0006 a p<0.10 Blacks: -0.0016  (3) Whites: -0.0006 Blacks: -0.0019  (4) Whites: -0.0006 Blacks: -0.0009 | Results shows the intervention impact of FSP on birth weight and LBW percentage, respectively in the white and black population.  They found this cash transfer improved birth outcomes, effects being larger in populations at greater  nutritional risk relative to Whites. |
| **Baker et al****(39)** | Earned Income Tax Credit | Birth weight in grams   LBW % | DD^1^ and DDD^1^ regression coefficient | **Birth weight (g)**  DD Model1^3^ 3.63 (p>0.10)  DD Model2 7.30 (p<0.01)  DDD Model2 13.8 (p<0.01)  **Low birth weight %**  DD Model1 -0.02 (p<0.01)  DD Model2 -0.028 (p<0.01)  DDD Model2 -0.0033 (p<0.01) | Results indicate that the 1993 expansion of EITC has been effective in increasing birth weights, though the magnitude of the effects is relatively small. The effect obtained with DDD model were more reliable- due to falsification test robustness- and they were higher in magnitude. |
| **Brownell et al****(24)** | Unconditional cash transfer | LBW rate  Small for Gestational Age rate� (SGA, weight below 10^th^ percentile) | Risk Ratio between who received the transfer and who did not | **LBW** 0.71 (95% CI 0.63-0.81)  **SGA** percentage 0.90 (95% CI 0.81-0.99) | The receipt of an unconditional  income supplement by very-low income  women during pregnancy  was associated with reduction in LBW and� Small for Gestational Age percentage, and an increase of Large for Gestational Age percentage |
| **Chung et al****(34)** | Universal unconditional cash transfer | Birth weight in grams   LBW % | DD^1^ regression coefficient | **Birth weight (g)**  Anticipatory^4^ 17.448 p<0.05  Realized 30.924 p<0.05  **LBW %**  Anticipatory -0.004 p<0.05  Realized -0.007 p<0.05 | They found that an additional $1,000 ($2,331 in 2011 dollars) increases birth weight by 17.7 g and decreased the likelihood of LBW. Though the APFD affects the overall population rather than just the poor, the effect was higher the income effect is higher for less educated mothers. |

|  |  |  |  |  |  |
| --- | --- | --- | --- | --- | --- |
| **Child outcome** | **Intervention** | **Outcome description** | **Measure of impact** | **Effect estimate** | **Results interpretation** |
| ***Birth weight*** |  | | | | |
| **Hoynes** **et al****(32)** | Earned Income Tax Credit | LBW % | DD^1^ regression coefficient | Parity 2+ versus Parity 1^5^: -0.35, *p<0.01*  Parity 3+ versus Parity 1: -0.53, *p<0.01*  Parity 3+ versus Parity 2: -0.34, *p<0.01*  Parity 2 versus Parity 1: -0.16, *p<0.05* | For single, low-education (12 years or less) mothers, a policy-induced treatment on the treated increase of $1000 in after-tax income is associated with a 0.17 to 0.31 percentage point decrease in low birth weight status. Given roughly 10.7 percent of treated children were low birth weight, this represents a 1.6 percent to 2.9 percent decline. |
| **Komro** **2019 et al****(27)** | Earned income Tax Credit | Birth weight in grams   LBW %  All results are stratified by race and shown according to the intervention characteristics^6^ | DD^1^ regression coefficient | Birth weight  *EITC, NR, < 10%*  Black:16.120, *p<0.01*  White:9.830, *p<0.01*  Hispanic:11.264, *p<0.01*  Non-hispanic:8.667, *p<0.05*  *EITC, R, <10%*  Black: 19.342, *p<0.01*  White:18.219, *p<0.05*  Hispanic:21.431, *p<0.01*  Non-hispanic:������� 15.530, *p<0.10*  �  *EITC, NR, 10%+*  Black:19.257, *p<0.01*  White:10.462, *p<0.01*  Hispanic:16.561, *p<0.01*  Non-hispanic:11.851, *p<0.01*  *EITC, R, 10%+*  Black:37.164, *p<0.01*  White:28.400, *p<0.01*  Hispanic:35.613, *p<0.10*  Non-hispanic:28.492, *p<0.01*  LBW %  *EITC, NR, < 10*%�  Black:-0.007, *p<0.01*  White:-0.002, *p<0.05*  Hispanic:-0.001, *p<0.10*  Non-hispanic:-0.004, *p<0.01*  *EITC, R, < 10%*��  Black:-0.009, *p<0.01*  White:-0.005, *p<0.01*  Hispanic:- 0.004, *p<0.05*  Non-hispanic:-0.006, *p<0.05*  *EITC NR, 10%+*�  Black:-0.006, *p<0.05*  White:-0.002, *p<0.01*  Hispanic:-0.002, *p<0.05*  Non-hispanic:-0.003, *p<0.05*  *EITC R, 10%+*  Black:-0.014, *p<0.01*  White:-0.007, *p<0.01*  Hispanic:-0.007, *p<0.01*  Non-hispanic:-0.009, *p<0.01* |  |
| **Komro** **2016 et al****(37)** | Minimum wage salary | LBW percentage | DD^1^ regression coefficient | **Crude^7^**: -1.9 (95% CI -3.1-0.7)  **Crude lagged**^8^: -2.2 (95%CI -3,6-0.8)  **Adjusted**: -1.1 (95% CI -2.1;-0.1)  **Adjusted lagged**: -1.3 (95%CI -2,7;0) | Results showed that increased state minimum wages are associated with reduced low birth weight births. The analyses were at the state level and refers to one dollar increase in minimum wage salaries |
| **Rosenthal et al****(36)** | Conditional Cash Transfer | LBW rate | Logistic regression coefficient (Odd Ratio) obtained through Instrumental Variable analysis | 0.53 (95%CI:� 0.23; -1.18) | Despite non-significance, the magnitude of effect is similar to the one obtained through ordinary logistic regression in which the exposure (i.e. intervention enrolment) is the one as reported in the dataset. |
| **Strully** **et al****(28)** | Earned Income Tax Credit | Birth weight in grams | DD^1^ regression coefficient | 15.704 (p<0.01) | Using a natural experimental strategy, they found that state EITCs increase birth weights in low income populations. Since they used dichotomous indicators for larger and smaller maximum state  credits, they found no evidence of a dose-response effect on birth weight |
| **Wehby** **et al****(38)** | Minimum Wage Salary | Birth weight in grams   LBW % | DD^1^ regression coefficient | **Birth weight (g)**  1.04 p<0.01  **LBW%**  �-0.00090 p<0.05 | The effect of the increase of 1$ in minimum wage is the increase of 4.04 grams in birthweight and a reduction of 0.009 % in rate of LBW.  They estimate �affected hours� as average annual working hours so that they could estimate the effect of 1000$ in annual income. Results of estimate are an increase of 8.468 grams and a reduction in 0.002% in LBW rate, both non-significant at 95% level |
| **Wicks-Lim et al****(33)** | Earned Income Tax Credit | LBW rate | DD^1^ regression coefficient | **Crude results**  OLS^9^ model -0.021 (SE 0.008) p<0.05  GLM model -0.331 (SE 0.162) p<0.05   **Adjusted results**  OLS model -0.022 (0.008) p<0.01  GLM model -0.330 (0.173) p<0.1 | NYS and NYC EITC expansions between 1997 and 2010 are linked to improved low birthweight rates in the city�s low-income  neighbourhoods. The magnitude of the EITC�s impact on low birthweight rates suggests ecological effects, and an  additional channel through which anti-poverty measures can serve as public health interventions |
| ***Child mental health*** |  | | | | |
| **Hamad et al 2016****(31)** | Earned Income Tax Credit | BPI score^10^.  The score was measured after 2 and 4 years^11^ | Linear regression coefficient (BPI score difference) | **2 year** **difference**  -0.46 (95%C.I: -0.86;-0.091)   **4 year** **difference**  -0.41 (95% C.I.: -0.91;-0.001) | Results suggest that there were positive effects on children�s behavioural problems in the sample overall. The effect magnitudes were approximately 5% of a standard deviation for every $1,000 of income.  Although these associations were modest, it is possible that persistent increases in income might bring about greater cumulative changes in child development |
| **Milligan et al****(26)** | Unconditional cash transfer | Social and motor development score^12^  Physical aggression score  Separation anxiety score  Indirect aggression score  Anxiety score | Regression coefficient | Social/motor development 0.187, *p<0.01*  Physical aggression -0.208, *p<0.01*  Separation anxiety -0.138  Indirect aggression -0.145  Anxiety -0.204, *p<0.10* |  |

1. As reported by the authors in the
   papers.

1. Estimated through Difference in
   Difference (DD) and/or Triple Difference (DDD)
2. For each outcome, they report estimates
   from four specifications with different controls (1) includes county and
   time (year x quarter) fixed effects, county per capita income, REIS
   county-level per-capita transfers, and 1960 county characteristics
   interacted with linear time (2) with state specific linear time trends,
   (3) with unrestricted state by year fixed effects, and (4) with county
   specific linear time trends
3. Since EITC 1993 expansion increased both
   the maximum credit for all families and generated significant differences
   in benefits for families with 2 or more children and families with 1
   child, model 1 uses mothers giving birth to their first or second child as
   a control (partial EITC reform effect) and model 2 uses as control mothers
   giving birth to their first child (full EITC reform effect)
4. The intervention group was divided in
   two periods, Anticipatory- born prior the intervention delivery but when
   they were expected to receive it, and Realized- born after individuals
   received the credit
5. The study used different models
   comparing parity of the mothers because EITC treatment corresponds to the
   number of children prior to the current birth. Model 1 uses women at second
   or higher pregnancy as intervention group and women at first as control.
   Model 2 uses separately women at second and third or higher pregnancy as
   control vs first pregnancy. Model 3 uses mother at third or higher
   pregnancy vs mother at second.
6. Intervention categories are defined in
   this way: NR/R (Non-refundable or Refundable amount) and intervention
   benefit size groups (reported as percentage of the federal amount, i.e. less than 10%, 10% or more). All EITC categories
   are compared with states with no EITC (reference).
7. Crude results are adjusted for state and
   year fixed effect only while adjusted results take in account also of
   race, poverty, cigarette sales and maternal age
8. Lagged results refers to 12 months after
   policy change
9. Two different models were used: Ordinary
   Least Square (OLS) regression and Generalised Linear Models (GLM)
10. 6 Different scales are used as Behavior Problems Index, Behavior
    Problems Scale, Survey Diagnostic Instrument Conduct Disorder. Result are reported as standard deviation of the scores,
    normalized
11. In the results BPI score difference was
    calculated both as 4 years after the intervention vs actual difference and
    2 years after the intervention vs actual difference
12. Scales used in this study are Canadian
    scales of parents reports developed for use in
    the National Longitudinal Survey of Children and Youth. Results are
    stratified per age. We took in account the impact on pre-schoolers at 36
    months of follow-up
